# The clinical impact of maternal COVID-19 on mothers, their infants, and placentas with an analysis of vertical transfer of maternal SARS-CoV-2-specific IgG antibodies

**DOI:** 10.1101/2022.02.01.22270179

**Authors:** JD Ward, C Cornaby, T Kato, RC Gilmore, D Bunch, MB Miller, RC Boucher, JL Schmitz, FA Askin, LR Scanga

**Author notes:** **Corresponding authors: Jeremy D. Ward** (*Address:* Department of Pathology and Laboratory Medicine, Campus Box #7525, Brinkhous-Bullitt Building, Chapel Hill, NC 27599-7525, United States; *Email*, **Lori R. Scanga** (*Address:* Department of Surgical Pathology, Women’s and Children’s Hospitals, 3rd Floor, Room 30149, 101 Manning Drive, Chapel Hill, NC 27514; *Email*).

## Abstract

**Introduction:** The effect of SARS-CoV-2 infection on pregnant mothers, the placenta, and infants is not fully understood and sufficiently characterized.

**Methods:** We performed a retrospective, observational cohort study in Chapel Hill, NC of 115 mothers with SARS-CoV-2 and singleton pregnancies from December 1, 2019 to May 31, 2021. We performed a chart review to document the infants’ weight, length, head circumference, survival, congenital abnormalities, and hearing loss, maternal complications, and placental pathology classified by the Amsterdam criteria.

**Results:** The average infant weight, length, and head circumference were all within the normal range for gestational age, the infants had no identifiable congenital abnormalities, and all infants and mothers survived. Only one infant (0.870%) became infected with SARS-CoV-2. Moderate and severe maternal COVID-19 were associated with increased caesarean section, premature delivery, infant NICU admission, and maternal respiratory failure, and were more likely in Type 1 (p=0.0055) and Type 2 (p=0.0285) diabetic mothers. Most placentas (n=63, 54.8%) showed normal or non-specific findings, while a subset had mild maternal vascular malperfusion (n=26, 22.6%) and/or mild microscopic ascending intrauterine infection (n=28, 24.3%).

**Discussion:** Most mothers with SARS-CoV-2 and their infants had a routine clinical course. Maternal SARS-CoV-2 infection was not associated with intrauterine fetal demise, infant death, congenital abnormalities, or hearing loss. Infant infection with SARS-CoV-2 was rare and not via the placenta. Most placentas had non-specific findings and a subset showed mild maternal vascular malperfusion and/or mild microscopic ascending intrauterine infection, which were not associated with maternal COVID-19 severity.

## 1. Introduction

SARS-CoV-2 is a novel enveloped single positive stranded RNA virus of the coronavirus family [1] that has infected hundreds of millions of individuals resulting in over 5 million deaths worldwide. This study was initiated at the beginning of the SARS-CoV-2 pandemic when little was known about the effects in pregnancy. Therefore, we wanted to determine if SARS-CoV-2 infection causes any specific histologic change in the placenta, the incidence of SARS-CoV-2 transmission to infants, and the survival outcomes of the mothers and their infants. Since the initiation of our study, there have been increasing reports on the effects of SARS-CoV-2 on the placenta. In a recent meta-analysis of these placental studies [2], no histological findings in the placenta were specific for COVID-19. Moreover, all studies to date have shown the incidence of trans-placental viral transmission to be very rare and when there is SARS-CoV-2 infection of the placenta, no specific viral changes have been noted [2]. However, a recent small study [3] demonstrated that placentas positive for SARS-CoV-2 via RNA ISH demonstrated chronic histocytic intervillositis, perivillous fibrin deposition, and trophoblastic necrosis. Interestingly, 63 percent of the infants in this study [3] tested negative for SARS-CoV-2, meaning that the placenta may be infected while the infant is not in a number of cases. Finally, there has been some speculation that moderate and severe maternal hypoxia may lead to maternal vascular malperfusion (MVM) changes [4]. Therefore, additional studies of SARS-CoV-2 infected mothers are needed to determine the common and rare effects of SARS-CoV-2 on the placenta.

To date, many of the studies on SARS-CoV-2 and its effect on the mother, infant, and placenta [4,5] have been in mothers infected in the third trimester. Due to the length of time for our study we were able to include a significant proportion of mothers who contracted SARS-CoV-2 in their first or second trimester. Moreover, we were able to follow these infants, in some cases over a year, for congenital birth defects and/or hearing impairment that may be associated with maternal SARS-CoV-2.

In our study, the incidence of maternal transmission was very low (0.870%) and was not via the placenta. After finding that only one infant in our original cohort became infected with SARS-CoV-2, we conscripted a second small cohort and mothers and their infants to determine if mothers infected with SARS-CoV-2 developed SARS-CoV-2-specific IgG antibodies and if these IgG antibodies were transferred to their newborns. We found that nearly all mothers, except for a single individual mother who was likely immunosuppressed from very recent bacterial sepsis, developed SARS-CoV-2-specifc IgG antibodies and transferred all or a majority of these antibodies to their infants. These findings are in keeping with several recent studies [6–8] that demonstrated mothers infected with SARS-CoV-2 do generate SARS-CoV-2-specific IgG antibodies and transfer them to their infants; however, these studies differ on the efficiency of the transfer. Our study adds to these data since we found that at least one of six SARS-CoV-2-specifc IgG antibodies we tested were transferred and retained by the infant and that this was irrespective of the trimester of the maternal SARS-CoV-2 infection.

Overall, our study found most mothers with SARS-CoV-2 infections delivered at term and their infants had medically insignificant to no complications. None of the infants or mothers died during the period of our study, which was over a year for many mother and infant dyads. We also found that no infants had any identifiable birth defects linked to maternal COVID-19 infection. Conversely, 13% of the mothers in our cohort who developed moderate or severe COVID-19, had a statistically significant increase in the incidence of premature births, delivery by C-section, maternal respiratory failure, maternal supplemental oxygen requirement, delivery complications, preeclampsia and admission of their infants to the NICU. Finally, we found that pre-existing diabetes mellitus was the most significant risk factor for developing moderate or severe maternal COVID-19.

## 2. Materials and methods

### 2.1. Study design

This was a retrospective observational consecutive cohort study of mothers with a positive SARS-CoV-2 PCR result during their current pregnancy who delivered at The University of North Carolina (UNC) Hospitals in Chapel Hill, NC, between December 1, 2019 and May 31, 2021, and whose placentas were evaluated by Anatomic Pathology. Their infants were included in the cohort. We excluded mothers and their infants who were not the product of a singleton birth and infants found to have genetic abnormalities. Data was obtained from the electronic medical record (EMR) in accordance with the University of North Carolina at Chapel Hill Internal Review Board-approved study parameters (IRB# 20-2944).

### 2.2. COVID-19 severity quantification

We utilized a simplification of the WHO ordinal scale [9] to categorize the maternal COVID-19 severity as asymptomatic, mild, moderate, or severe with the numerical score of 1, 2, 3, 4, respectively.

### 2.3. Newborn weight, length, and head circumference percentiles

Newborn weight, length, and head circumference percentiles were based on the WHO 0-2-year-old growth charts [10].

### 2.4. SARS-CoV-2 serologic testing

In a smaller non-consecutive study, from July 10, 2021 to August 31, 2021, we identified 10 mothers who had a positive SARS-CoV-2 PCR during their current pregnancy, had not received a COVID-19 vaccination, delivered their infants at UNC Hospital main campus, and both the mother and their infant had enough blood remaining in the laboratory to perform SARS-CoV-2-specific antibody assays. SARS-CoV-2-specific IgG antibodies against the spike tetramer, S1, RBD, S2, and nucleocapsid proteins were detected from patient serum or plasma utilizing the LabScreen COVID plus multiplex bead assay (OneLambda) per manufacturer instructions on the LabScan3D platform (Luminex). These same samples, in addition to appropriate positive and negative controls, were tested for SARS-CoV-2 neutralizing antibodies using a surrogate neutralizing antibody assay (GeneScript) per the manufacturer’s instructions.

### 2.5. Placenta processing and examination

The placentas were processed and evaluated using standard methods [11]. All noted gross and histologic findings were quantified from the final pathology reports in the EMR. The placentas were assigned to one (or more) of the five Amsterdam consensus group criteria or classified as normal/non-specific if the placental findings did not meet the threshold for one of these classifications [12].

### 2.6. Immunohistochemical and RNAScope analysis

Immunohistochemistry for SARS coronavirus nucleocapsid (Invitrogen PA1-41098) and RNA in situ hybridization for SARS-CoV-2 RNA (Advanced Cell Diagnostics, 848561) for selected placental tissues were performed as previously described including appropriate positive and negative controls [13,14].

### 2.7. Statistical analysis

All statistical analyses were performed using GraphPad Prism version 9.2.0 except for polynomial and linear trend analysis, which were performed using Microsoft Excel (Microsoft 365). Chi-square for linear trend, Fischer exact test, Mann-Whitney test, or one-way ANOVA with multiple comparisons between groups using Tukey’s test were used as appropriate. Survival curve comparison was performed using the Gehan-Breslow-Wilcoxon test. P values <0.050 were considered significant.

## 3. Results

### 3.1. Maternal Characteristics

One hundred twenty-five mothers were identified, and 115 mothers, including their placentas and infants, were included in the study (Figure 1). Clinical information that was collected and analyzed is listed in Table 1. The mean (median) maternal age was 29 (28) years with a range of 15 to 42 years old. 58% of the mothers were Hispanic, which is substantially increased compared to 9.6%, the estimated Hispanic statewide population of North Carolina [15]. Nine mothers (7.83%) were infected with SARS-CoV-2 in their first trimester, 27 mothers (23.5%) were infected in their second trimester, with the remaining 79 mothers (68.7%) infected in their third trimester. The mean (median) of the first positive test result was 30.5 (34.6) weeks gestation, and ∼35% tested positive on admission. The average (median) gestational age at delivery was 38.3 (39 1/7) weeks and 69% were vaginal and 31% were via C-section. 85.2% of mothers were asymptomatic (n=37) or had mild (n=61) symptoms, 13.0% of mothers had moderate (n=9) or severe (n=6) COVID-19, and 1.74% (n=2) mothers did not have their symptoms reported.

**Table 1:**
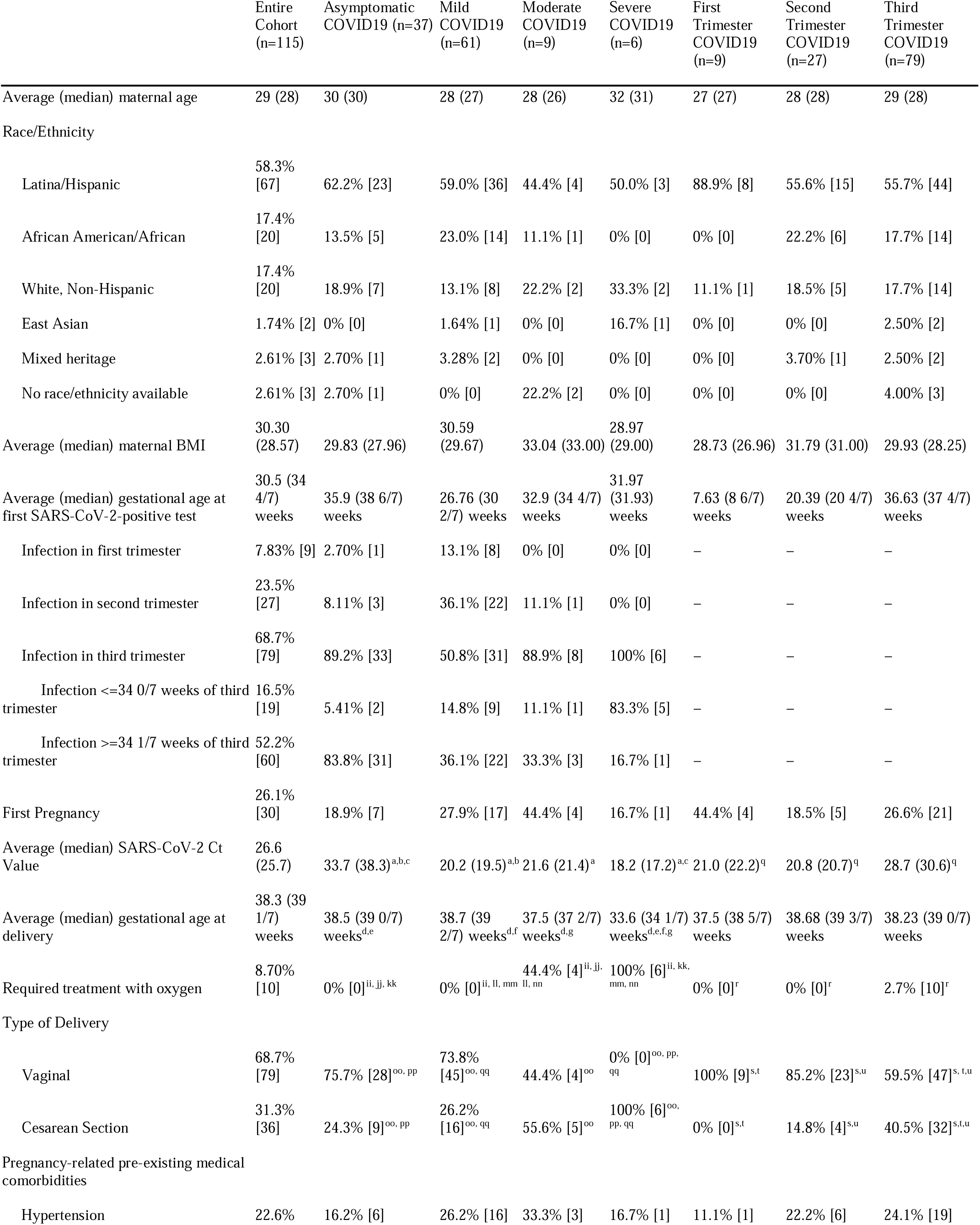
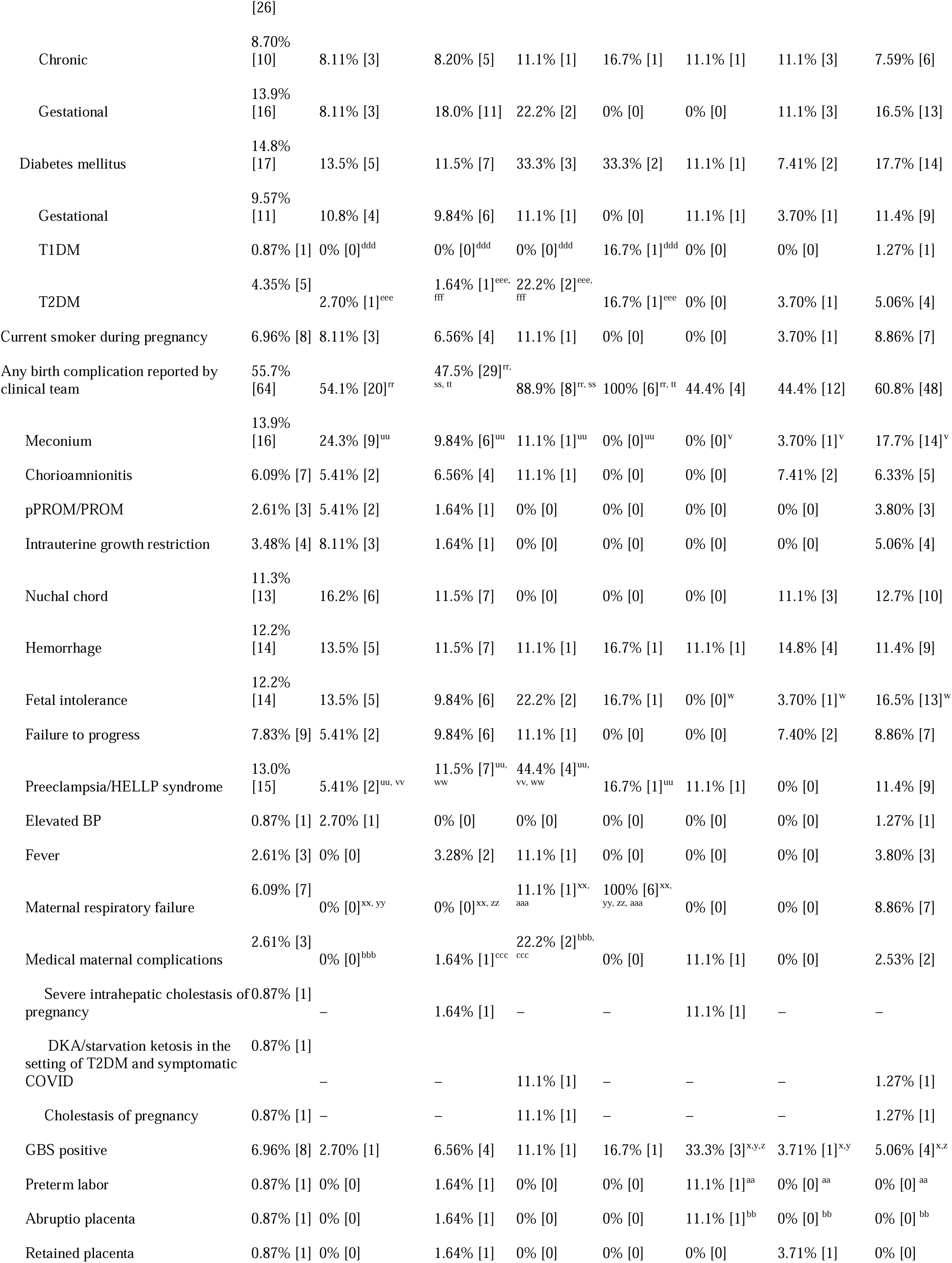
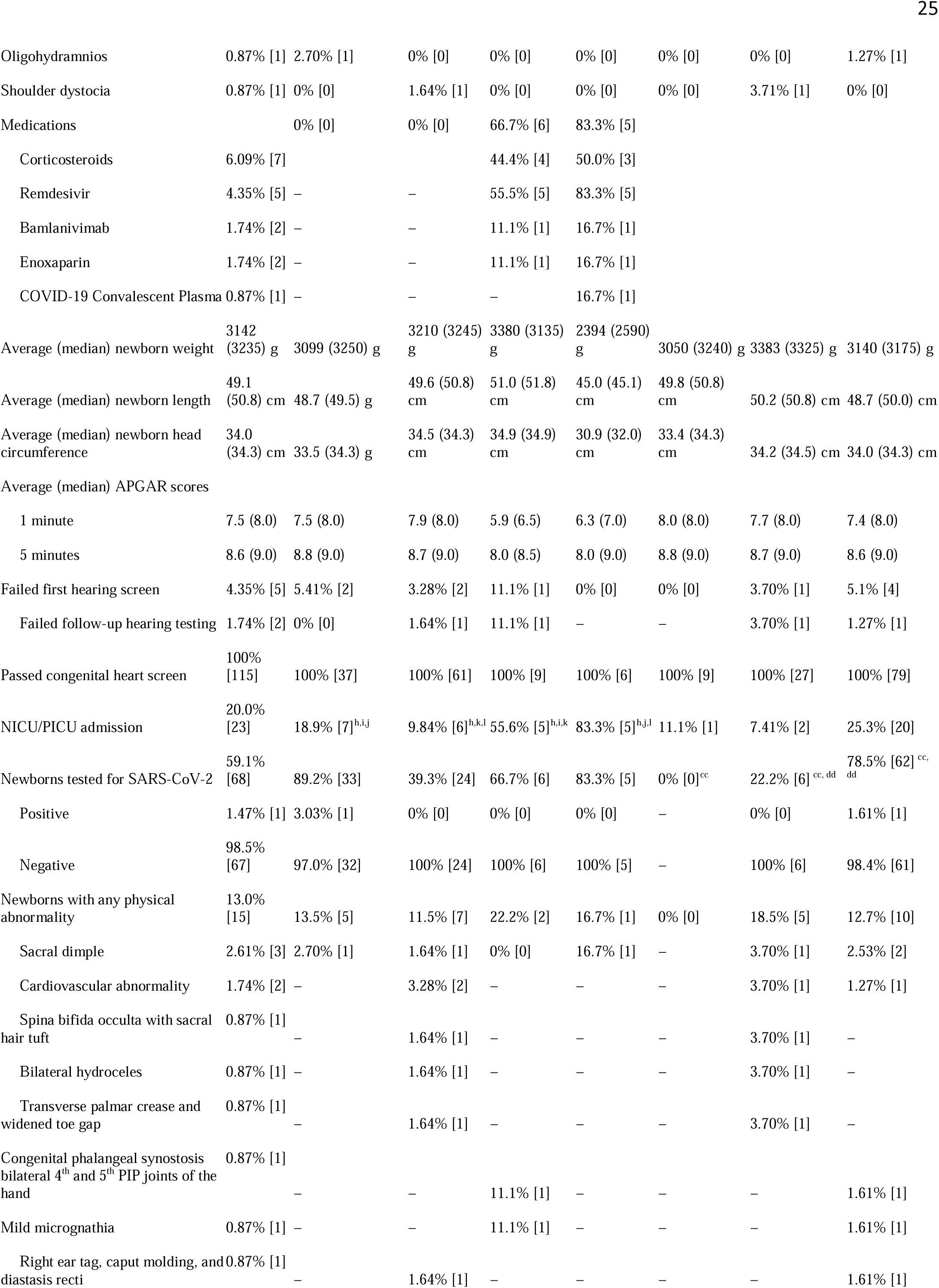
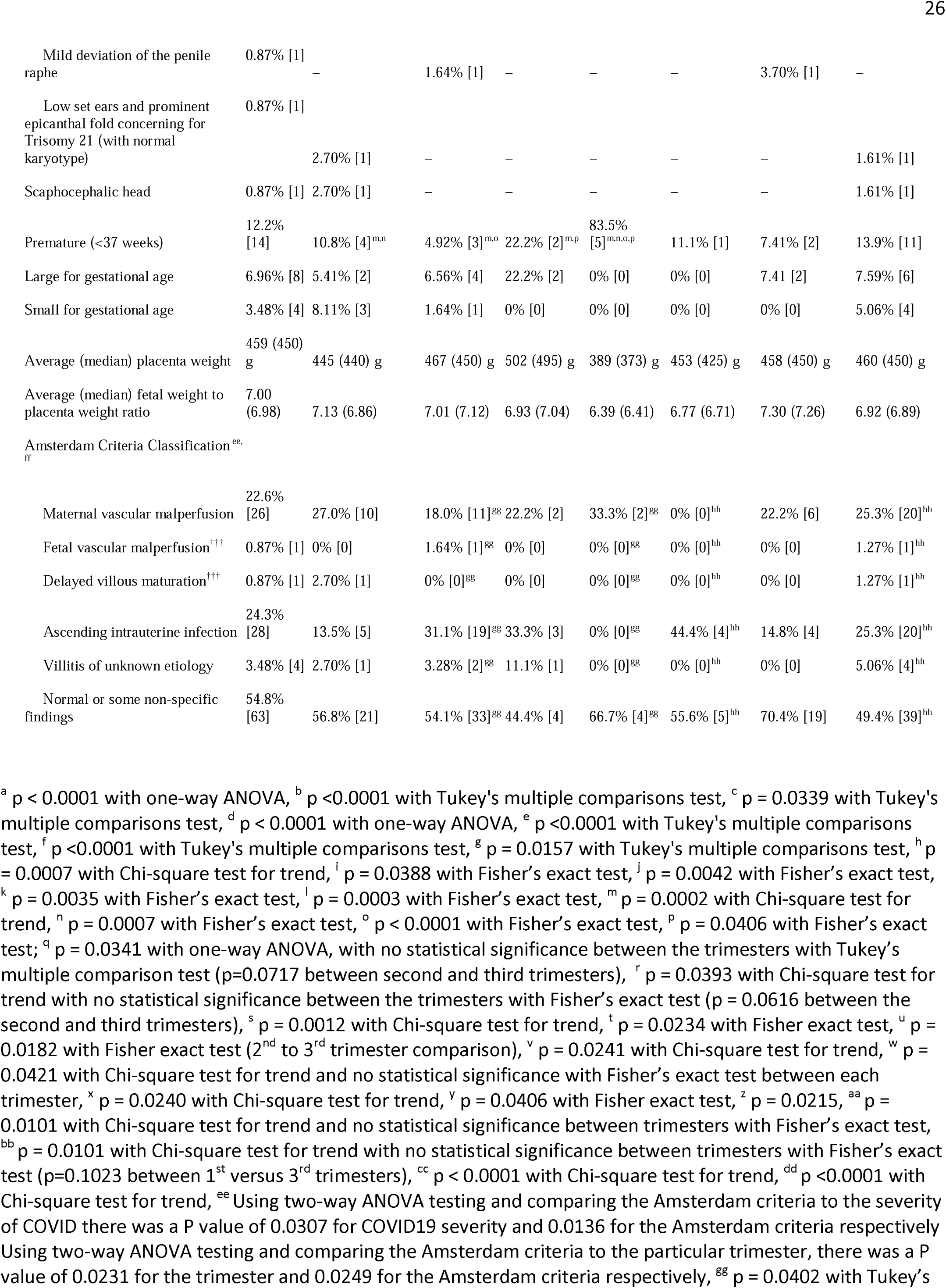
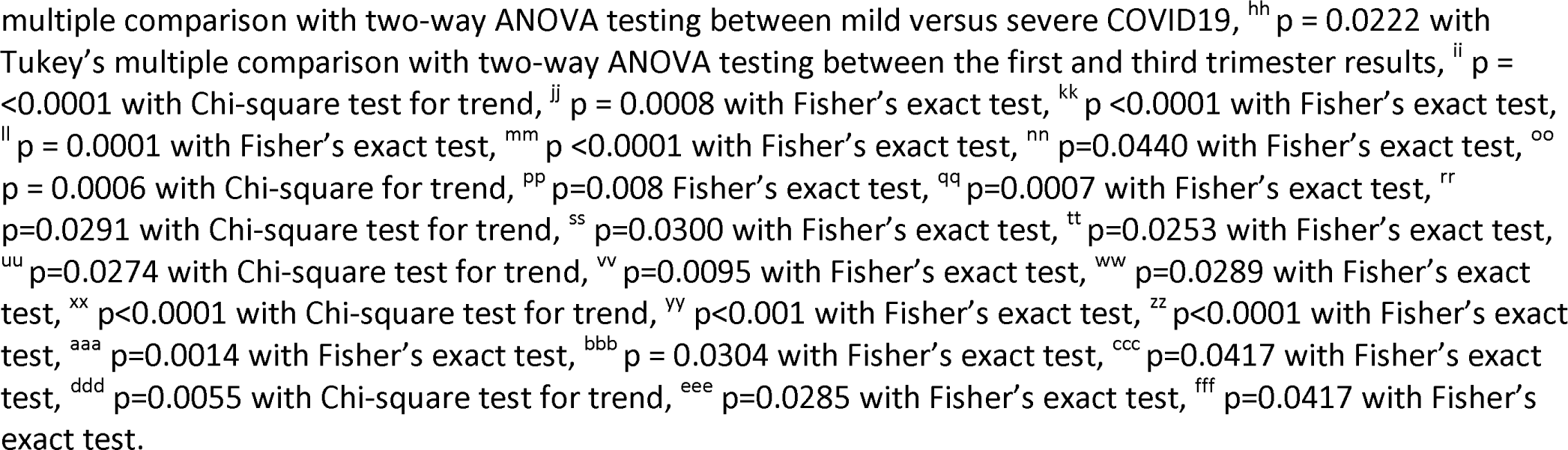

**Figure 1:**
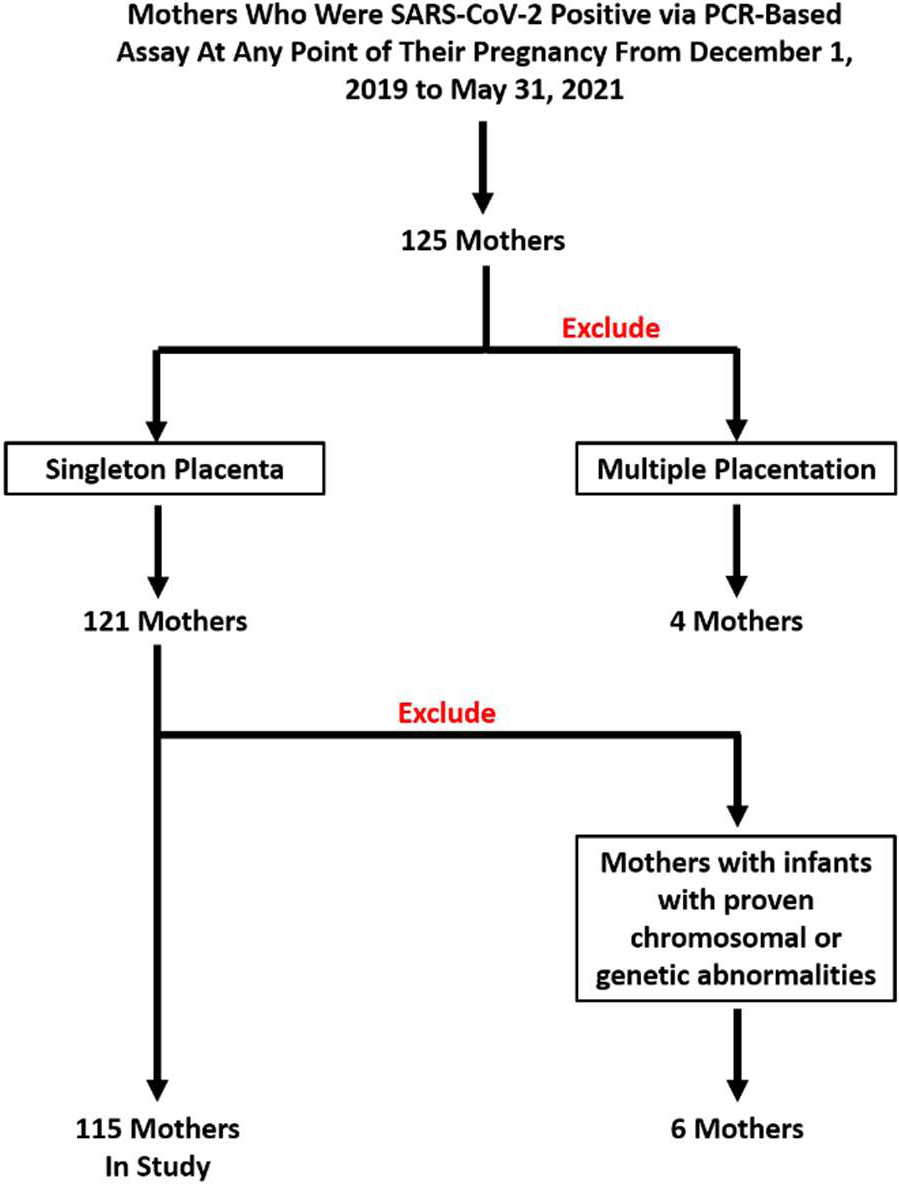
Flow diagram of selection of study cohort.

### 3.2. Severity of Maternal COVID-19 Infection

Mothers with severe or moderate COVID-19 were statistically more likely to be Type 1 (p=0.0055) or Type 2 (p=0.0285) diabetics, have respiratory failure (p<0.0001), require supplemental oxygen (p<0.0001), deliver via C-section (p=0.0006), have any delivery complication (p=0.0291), preeclampsia (p=0.0274), or premature infants (p=0.0002) compared to mothers who were mildly ill (less likely) or asymptomatic (least likely). Three of the six severely ill mothers delivered at 26-32 weeks and one severely ill and two of the nine moderately ill mothers delivered at 35-36 weeks due to respiratory failure. 8.70 percent of mothers required supplemental oxygen, which included 44.4% of the moderately and 100% of the severely ill mothers (p<0.0001). Similarly, maternal respiratory failure attributed to COVID-19 was seen in seven mothers (6.09%) and was significantly increased (p<0.0001) in the moderately (11.1%) and severely ill (100%). There was a strong tendency for increased maternal medical complications (p=0.0668) from asymptomatic (least likely) to severely ill (most likely). Mothers with severe COVID-19 delivered prematurely, at an average (median) gestational age of 33.6 (34 1/7) weeks compared to 38.5 (39 0/7) weeks with asymptomatic infection (p<0.0001), 38.7 (39 2/7) weeks with mild COVID-19 (p<0.0001), and 37.5 (37 2/7) weeks with moderate COVID-19 (p=0.0157). Conversely, there was a statistically significant inverse correlation for clinically detected meconium (p=0.0457) in asymptomatic mothers (most likely) versus severely ill (least likely) mothers.

Subgroup analysis demonstrated a statistically significant difference in delivery complications between mild versus moderate (p=0.0300) and mild versus severe (p=0.0253) maternal infections. There was a strong tendency for increased delivery complications between asymptomatic versus moderate (p=0.0691) and asymptomatic versus severely ill (p=0.0662) mothers. There was no statistically significant difference in delivery complications between moderate versus severe maternal infections (p>0.9999).

### 3.3. Trimester of Maternal COVID-19 Infection

There was a statistically significant increase in the maternal requirement for supplemental oxygen (p=0.0393), incidence of C-section (p=0.0012), presence of clinically noted meconium (p=0.0241), and fetal intolerance (p=0.042) when progressing from maternal SARS-CoV-2 infection in the first trimester (least risk) to the third trimester (most risk). There was a statistically significant increased odds ratio (OR 3.91, 1.31-11.2, 95% CI) of C-section with a third trimester SARS-CoV-2 infection versus a first trimester (p=0.0234) or a second trimester (p=0.0182) maternal infection. Conversely, there was a statistically significant increase in group B *Streptococcus* (GBS) infection (p=0.0240), preterm labor (p=0.0101), and *abruptio placentae* (p=0.0101) from first trimester (most risk) to third trimester (least risk) maternal infection.

### 3.4. Infant Characteristics

In the cohort, there were slightly more male infants than female infants (Figure 2A). Infant birthweights (Figure 2B), newborn lengths (Figure 2C), and head circumferences (Figure 2D) were all within normal parameters for gestational age. There was not a statistically significant difference between the male and female weights (p=0.2273) or the male and female infant head circumferences (p= 0.5635). There was a tendency for the female infants to be shorter than the male infants (p=0.0680). On subgroup analysis, there was a statistically significant decrease in the mean infant weights with severe versus mild (p=0.0246) or moderate (p=0.0286) maternal COVID-19 and a tendency toward lower birth weights (p=0.0675) with asymptomatic maternal infections. There was a statistically significant decrease in the mean infant length (p=0.0372) with moderate or severe maternal infections. Similarly, there was a statistically significant decrease in infant head circumferences with severe maternal infections versus mild infections (p=0.0428).

**Figure 2:**
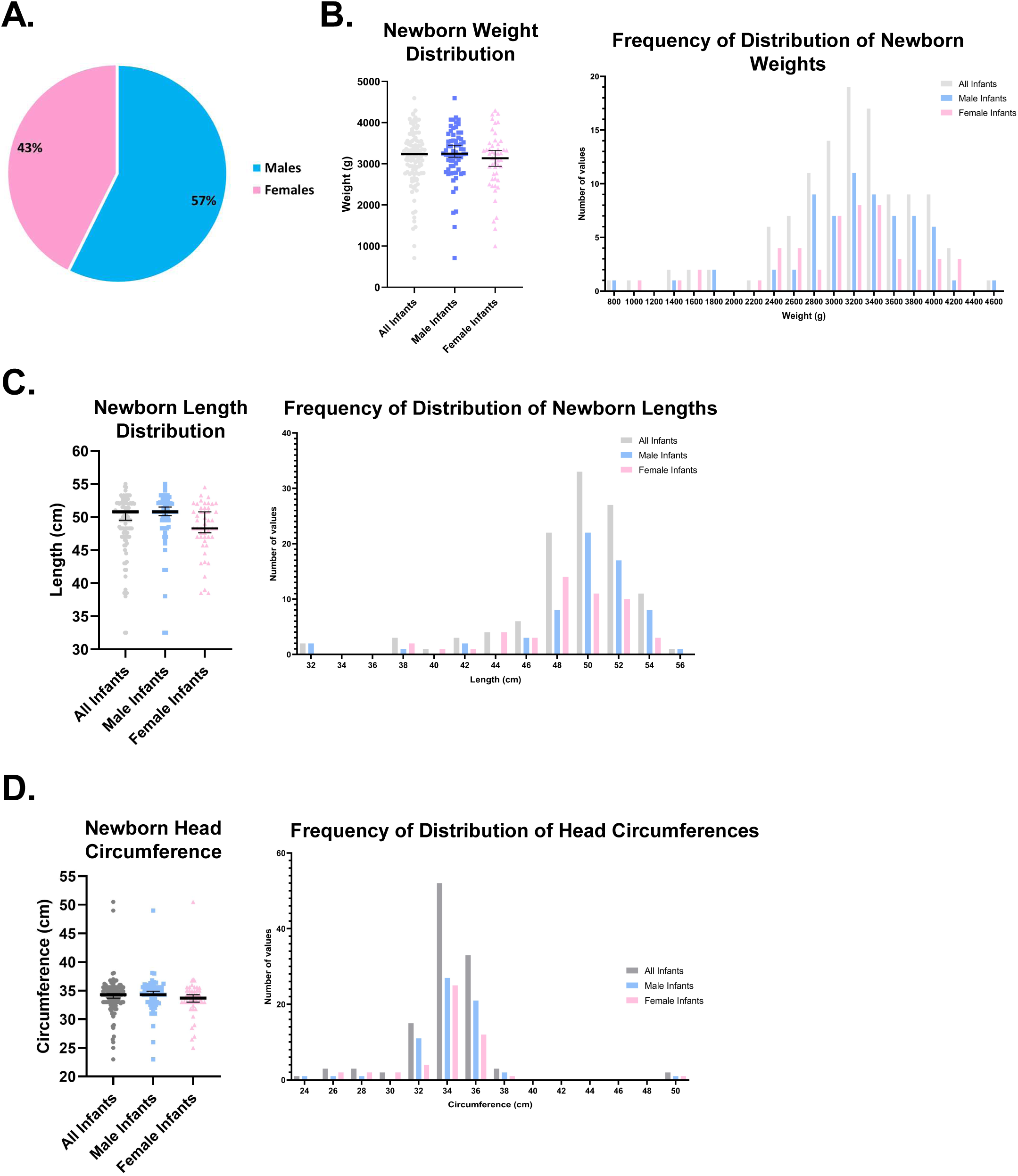
Sex, weight, length, and head circumference distribution of infants within the study cohort.

Twenty-three (20.0%) of the infants were admitted to the neonatal intensive care unit (NICU) after birth. A severe maternal infection was associated with an increased incidence of admission to the NICU (p=0.0007) and premature birth (p=0.0002). Subgroup analysis showed significantly increased admission to the NICU after moderate (p=0.0388) or severe (p=0.0042) versus asymptomatic maternal infections. Similarly, mild versus moderate (p=0.0035) and mild versus severe (p=0.0003) maternal COVID-19 were associated with an increased risk of NICU admission. Asymptomatic versus severe (p=0.0007) and mild versus severe (p<0.0001) maternal COVID-19 were associated with a statistically increased risk of premature birth. Finally, all infants were alive at the completion of the study on May 31, 2021 and there were no incidents of fetal demise (Supplemental Figure 1).

Fifteen (13.0%) infants had a reported physical abnormality. The most common (Table 1) was a deep sacral dimple in three infants (2.61%). However, there were no major or recurring physical abnormalities seen in any of the infants, including those born after a first trimester maternal SARS-CoV-2 infection. Five (4.3%) infants failed their newborn hearing screen. Of these 5 infants, 3 passed on follow up hearing tests, and 2 infants failed again (1.7%). There is insufficient follow up in these two infants as to the cause of hearing loss and there was no indication that it was secondary to maternal SARS-CoV-2 infection. All infants were screened for congenital heart disease via pulse oximetry and 100% passed. On follow-up, there were 2 infants that were found to have completely dissimilar cardiovascular abnormalities.

### 3.5. Maternal Transmission

Sixty-eight infants (59.1%) were tested for SARS-CoV-2 RNA at 24 hours of life, with many of the infants (n=40) receiving a second test at 48 hours of life. Of these 68 infants, only one infant (1.47%) was found to be positive for SARS-CoV-2 by PCR at 24 hours. If we assume that the untested infants were also negative, the overall incidence of SARS-CoV-2 infection was 0.870% in our cohort. Many of the infants stayed in the same rooms with their mothers and nearly all the infants were breastfed. There was no evidence of placental SARS-CoV-2 infection, meaning that transmission of SARS-CoV-2 was likely not via the placenta (Figure 3 and Supplemental Figure 3). Therefore, we hypothesized that the lack of SARS-CoV-2 infection in these infants may be due to vertical transfer of SARS-CoV-2-specfic IgG antibodies. In a smaller non-consecutive study, from July 10, 2021 to August 31, 2021, we identified 10 mothers who had a positive SARS-CoV-2 PCR during their current pregnancy, had not received a COVID-19 vaccination, delivered their infants at UNC Hospital main campus, and both the mother and their infant had enough blood remaining in the laboratory to perform SARS-CoV-2-specific antibody assays. Nine of 10 mothers (90%) had SARS-CoV-2-specific IgG antibodies and these antibodies were detected in the blood of their infants. One mother (10%) did not develop a significant antibody response to SARS-CoV-2, and her infant tested positive for SARS-CoV-2 by PCR. Similar confirmatory results were found with a surrogate assay for SARS-CoV-2 neutralizing antibodies (Figure 4 and Supplemental Table 1).

**Figure 3:**
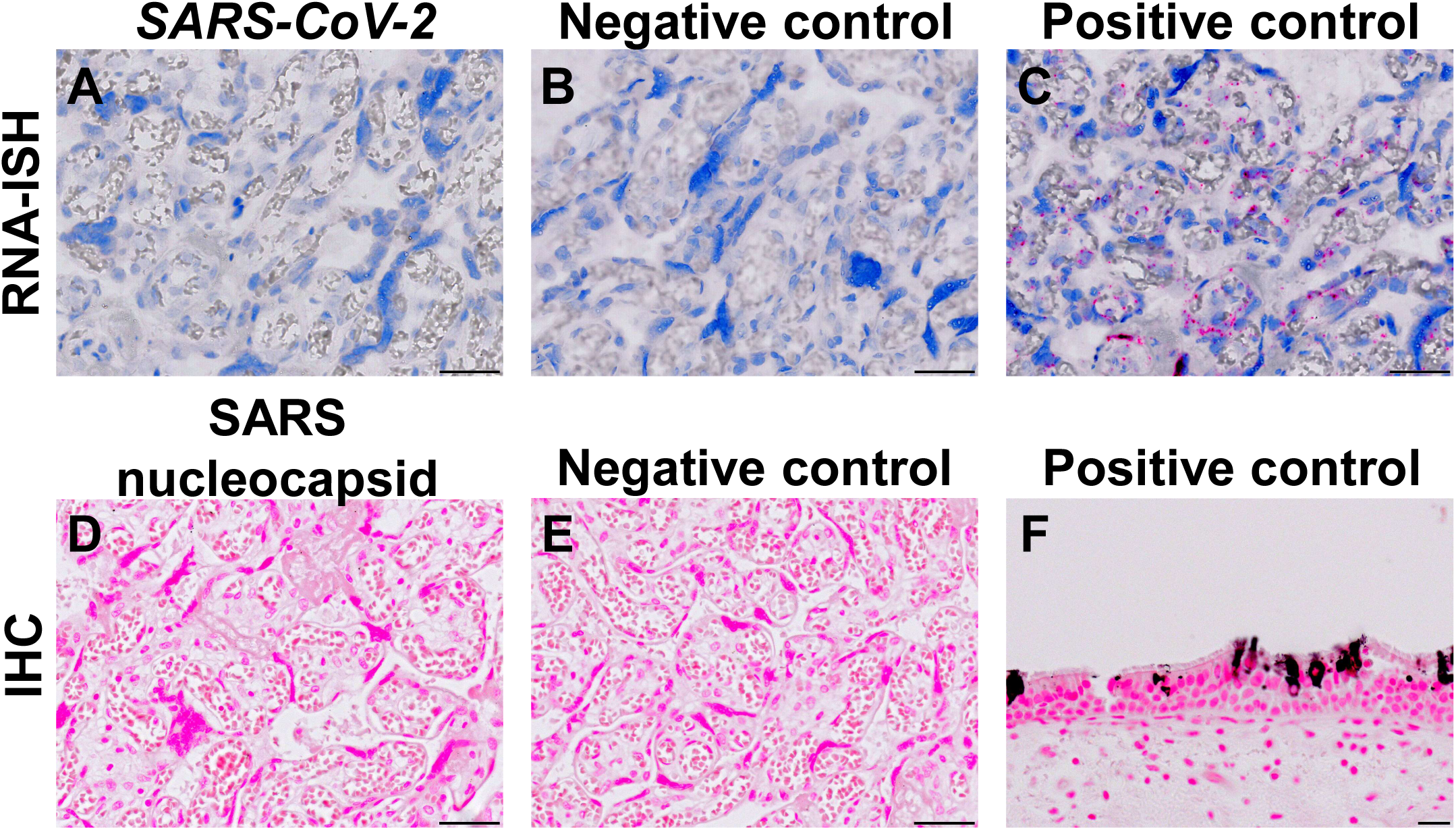
RNAScope for SARS-CoV-2 and immunohistochemistry for SARS-CoV-2 spike protein demonstrate the absence of placental infection with SARS-CoV-2 in the single SARS-CoV-2 positive infant in our cohort. (A-C) RNA-ISH of placenta for SARS-CoV-2 RNA (A) and negative (bacterial *DapB*) (B) and positive (*UBC*) (C) controls. (D-E) IHC of placenta for SARS nucleocapsid (D) and isotype control (E). (F) IHC of COVID-19 trachea as a positive control. Bars: (A-E): 50 μm. (F) 20 μm.

**Figure 4:**
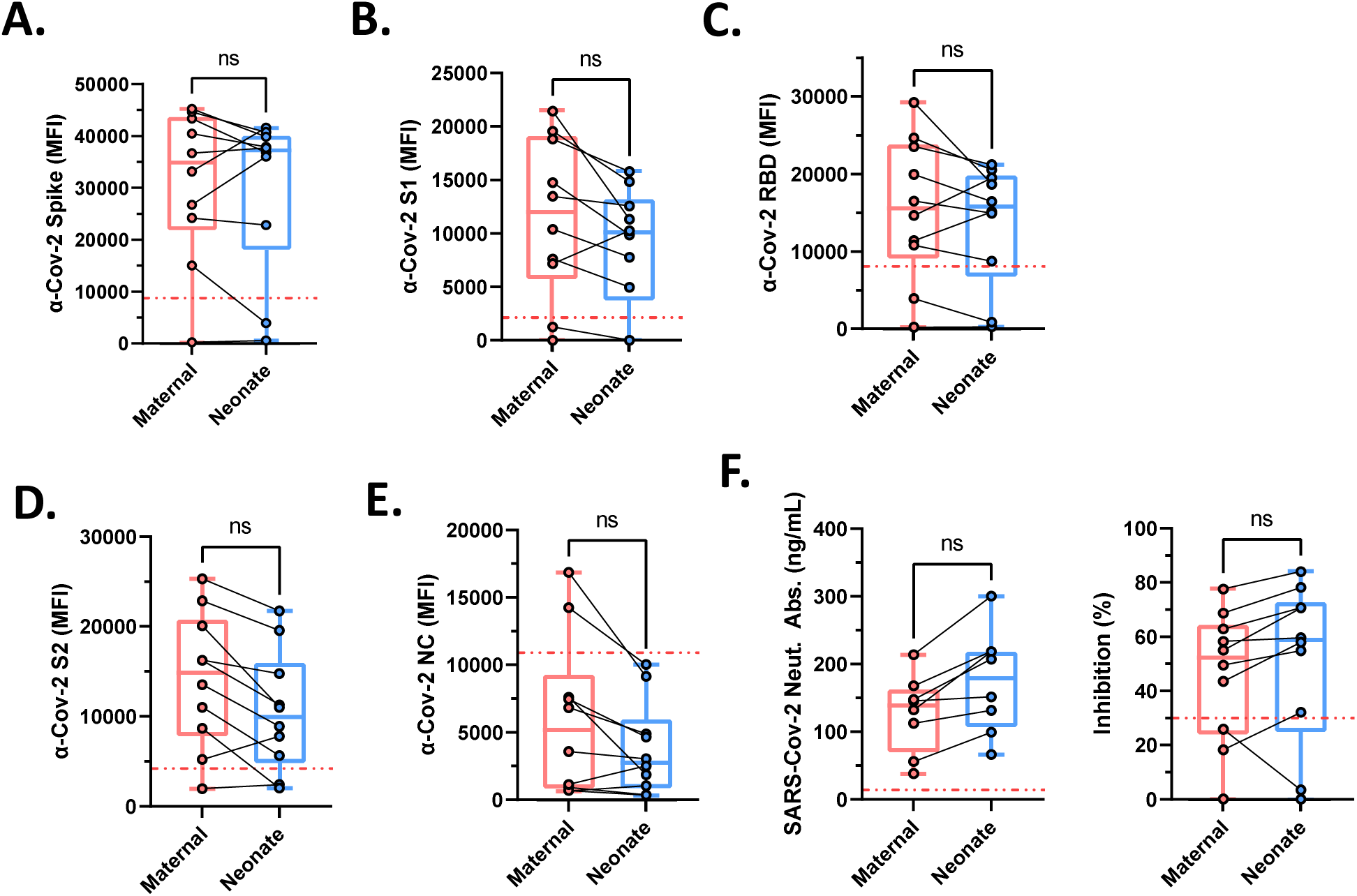
SARS-CoV-2-specific IgG antibodies in mothers and their infants. Box and whisker plots demonstrating **A:** SARS-CoV-2 spike IgG antibody, **B:** SARS-CoV-2 spike glycoprotein S1 subunit IgG antibody, **C:** SARS-CoV-2 receptor-binding domain IgG antibody, **D:** SARS-CoV-2 spike glycoprotein 2 subunit IgG antibody, and E: SARS-CoV-2 Nucleocapsid IgG antibody. F: SARS-CoV-2-surrogate neutralizing IgG antibody concentration and percent inhibition. The dashed red line is the threshold for a positive test with each respective antibody assay. ns = not significant. n=10.

### 3.6. Placental Pathology

54.8% of the placentas had normal or non-specific findings based on the Amsterdam criteria (Figure 5A; see Supplemental Figure 2 for the incidence of specific gross and microscopic findings) [12]. The remaining placentas demonstrated maternal vascular malperfusion (MVM, 22.6%), ascending intrauterine infection (AIUI, 24.3%), villitis of unknown etiology (3.48%), fetal vascular malperfusion (FVM, 0.870%), and/or delayed villous maturation (0.870%). There was no statistically significant difference in any of the Amsterdam criteria or normal/non-specific findings among the severity groups of maternal infection or from first to third trimester COVID-19 infection.

**Figure 5:**
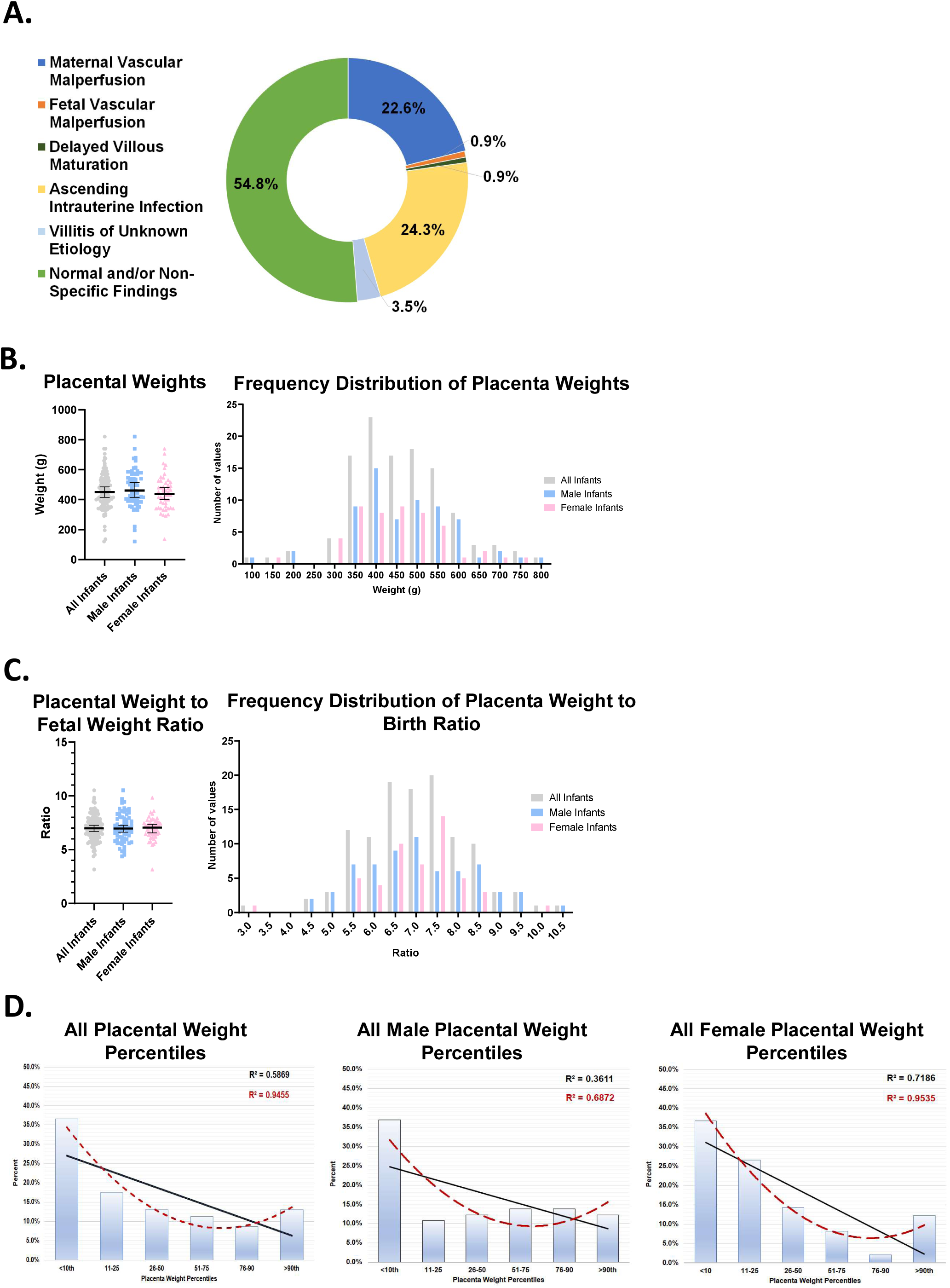
Amsterdam criteria, placental weight, placental weight to birth weight ratio, and placental weight percentiles of placentas within the study cohort. **A:** Amsterdam criteria classification of the evaluated placentas. Maternal vascular malperfusion: n=26, 22.6%; Fetal vascular malperfusion: n=1, 0.870%; Delayed villous maturation: n=1, 0.870%; Ascending intrauterine infection: n=28, 24.3%; Villitis of unknown etiology: n=4, 3.48%; Normal and/or non-specific findings: n=63, 54.8%. **B-C:** Scatter dot plot and bar graph of individual distribution of placental weights **(B)** and placental weight to birth weight ratio **(C)**. Solid line of scatter dot plot represents median with error bars representing 95% confidence intervals. **D:** Placental weight percentiles[25] for all placentas, male, and female placentas. Solid black line represents the linear trend line. Dashed red represents the 2^nd^ order polynomial trend line.

We performed a subgroup analysis of the placentas with MVM by eliminating cofounding causes, including preterm delivery (<37 0/7-weeks), maternal diabetes mellitus, hypertension, and/or pre-eclampsia. The incidence of MVM decreased from 22.6% (n=26) to 13.6% (n=9), which was not statistically significant (p=0.174). We also performed an analogous subgroup analysis of BMI (p=0.563), maternal age (p=0.769), vaginal birth versus C-section (p=0.171), and supplemental oxygen requirement (p=0.382), all of which were not statistically significant. Overall, none of the most common risk factors of MVM significantly explained the increase in MVM, suggesting that SARS-CoV-2 may be a contributing factor.

There was not a statistically significant difference (p=0.454) between the male and female placental weights (Figure 5B). The mean (median) placenta weight to birth weight ratio was 7.00 (6.98), 7.03 (6.96) for males, and 6.96 (7.04) for females, which was not statistically different (p=0.948) (Figure 5C). There was a trend of all the placenta weights toward lower percentiles, with 36.5% (36.9% of the males and 36.7% of the females) being less than the 10^th^ percentile (Figure 5D). The >10^th^ weight percentiles were largely normally distributed overall and in the male infants, while the female infants showed a greater skew of the placental weights to the lower percentiles.

## 4. Discussion

No significant physical or growth abnormalities were identified in any of the infants, including those from first or second trimester maternal COVID-19 infections. There was no fetal demise, and all the infants and mothers were alive at the end of the study. There was no evidence of infant hearing loss or cardiovascular abnormalities conclusively linked to maternal SARS-CoV-2 infection. There was only one incidence of SARS-CoV-2 transmission between a mother and infant (0.870%), for which we found no evidence of placental transmission. In a small sample (n=10), the majority (90%) of maternal SARS-CoV-2 infections generated maternal SARS-CoV-2-specific IgG antibodies that were transferred to their infants, likely accounting for the low transmission of SARS-CoV-2 to infants during pregnancy and puerperium. Mothers with moderate or severe maternal infections (13%) were statistically more likely to have preexisting Type 1 or Type 2 diabetes mellitus, and to have multiple and/or more severe maternal complications including respiratory failure, supplemental oxygen requirement, C-section, and preterm delivery. Twenty percent (23) of the infants in our study were admitted to the NICU, with most of these from mothers with moderate or severe COVID-19.

A strength of our study was its relatively large size and its duration, which allowed us to identify first and second trimester maternal SARS-CoV-2 infections and to follow maternal and infant outcomes. Another feature of the study was that most of the mothers (∼90%) in our cohort did not receive specific therapy for SARS-CoV-2, and none of the mothers were vaccinated, allowing our study to characterize the natural course of SARS-CoV-2 on the placenta, mother, and infant. However, a limitation of the study was the lack of truly normal control placentas. The placentas received for pathologic examination at UNC are ordered by clinicians for known or suspected maternal, placental, or fetal conditions and are not sent from uncomplicated deliveries. Our study also lacked blinding to the maternal diagnosis of SARS-CoV-2 infection and allowed access to the mothers’ chart, which may present a risk for bias. However, since our results were similar to previously published studies [4],[11,16], it is likely that this potential bias did not significantly affect the placental examination.

Recently, it has been shown [17–21] that maternal transmission of SARS-CoV-2 during pregnancy is a rare event, which was confirmed in our current study. A recent review [22] found that breastfeeding, mother-to-infant contact, and the method of delivery did not increase the risk for neonatal SARS-CoV-2 infection. A possible reason for this protection is vertical transfer of SARS-CoV-2-specfic antibodies from the mother to the infant. In support of this hypothesis, several recent studies [6–8] have demonstrated the vertical transfer of SARS-CoV-2-specific IgG antibodies. Interestingly, these studies report an efficiency of transfer of these antibodies as low as 25 percent [8] and as high as 87 percent [6]. However, these studies characterized the vertical transfer of antibodies targeted against a single SARS-CoV-2 epitope. Conversely, our study simultaneously characterized the transfer of 6 separate SARS-CoV-2-specific IgG antibodies with different epitopes in mother/infant dyads. While we found that not every individual antibody successfully transferred from the mother to the infant, when looking at the transfer at least one antibody from the mother to their infant, the overall efficiency of transfer was 100 percent. This difference in characterizing six separate SARS-CoV-2-specific antibodies may be why previous studies [7,8] reported a lower efficiency of placental transfer and why infants remained protected even when some of the SARS-CoV-2 antibodies were not transferred to them. In support of this hypothesis, the one mother in our study who did not develop an antibody response at all to SARS-CoV-2, her infant became infected with SARS-CoV-2. This suggests that the complete absence of generating maternal SARS-CoV-2-specific IgG antibodies increases the risk of infection of the infant by the mother. Therefore, testing mothers and/or infants for these SARS-CoV-2 IgG antibodies may be warranted and considered as a part of the discussion with a mother deciding whether to room in with their infant while infected with SARS-CoV-2. Nevertheless, additional studies on the transfer and duration of the protective function of these SARS-CoV-2-specific antibodies are needed.

Consistent with previous reports [2,4,11], over half of the placentas in our study had no significant gross or microscopic abnormalities. A minority of the placentas demonstrated MVM (22.6%) and/or AIUI (24.3%), and these placental findings were typically classified as mild. Notably, the average placental weight to newborn birth weight ratio was in the normal range, indicating the majority of placentas did not have significant functional abnormalities. This finding is consistent with most of the infants being of normal weight and size with no definitive abnormalities at birth. However, this study was conducted before the emergence of the delta and omicron variants in North Carolina. A recent report [23] and experience from our institution (unpublished data) preliminarily indicates that the delta variant may be associated with more adverse fetal outcomes and more specific placental pathology. Therefore, future studies will be needed to determine if there are differences in placental effects and maternal and fetal outcomes among the SARS-CoV-2 variants.

Fortunately, most mothers with SARS-CoV-2 infections acquired during pregnancy delivered at term and had uncomplicated vaginal deliveries. 31% of the deliveries in our cohort were by C-section, which is the national average for cesarean delivery in the United States [24]. However, moderate or severe COVID-19 maternal infection had an incidence of C-section of 55.6% (n=5) and 100% (n=6), respectively. In addition, pre-existing maternal diabetes mellitus was found to be a significant risk factor for developing moderate or severe maternal COVID-19 in our cohort, indicating that currently pregnant women or women wishing to become pregnant with diabetes mellitus should be encouraged to be vaccinated in order to prevent moderate to severe COVID-19 and its associated complications of delivery. Finally, we found that a subset of placentas demonstrated MVM with weights below the 10^th^ percentile, which was not linked to the severity of the maternal COVID-19 infection nor to known risk factors such as hypertension, preeclampsia, or maternal diabetes. Therefore, future studies will be needed to determine if there are specific circumstances (e.g., underlying genetic factors, specific immune responses, particular SARS-CoV-2 variants, etc.), along with the timing of the maternal SARS-CoV-2 infection that correlates with the development of MVM and significantly decreased placental weight.

## Data Availability

All data produced in the present study are available upon reasonable request to the authors.

## Disclosure of interests

The authors declare no conflicts of interest.

## Contribution to authorship

JDW, FAA, and LRS conceptualized and designed the study. JDW, CC, JLS, TK, RCG, RCB, DB, MBM, collected the data for the study. All authors contributed analysis and interpretation of data and to writing and/or editing of the manuscript and have seen the final version of the manuscript.

## Financial support

This research did not receive any specific grant from funding agencies in the public, commercial, or not-for-profit sectors.

## Acknowledgements

SARS-CoV-2 positive tracheal sections from a COVID-19 autopsy were provided by Dr. Alain Borczuk (Weill Cornell Medicine) under IRB-approved protocols # 20-04021880 and 20-04021796. We thank Shelby Currier, Nicky Gerken, Steve Holmes, April Kemper, and Andre Phelan who are pathology assistants at UNC for their gross dissection and histologic sampling of many of the placentas in this study. We thank the Obstetric, Family Medicine, and Pediatric nurses, residents and faculty for their hard work caring for the mothers and their infants in this study. Finally, we thank the mothers and infants involved in this study.

## Notes

### Competing Interest Statement

The authors have declared no competing interest.

### Funding Statement

This study did not receive any funding.

### Author Declarations

Ethics committee/IRB of the University of North Carolina gave ethical approval for this work. Specifically, data was obtained from the electronic medical record in accordance with the University of North Carolina at Chapel Hill Internal Review Board-approved study parameters (IRB# 20-2944).

## References

[1] P. V’kovski, A. Kratzel, S. Steiner, H. Stalder, V. Thiel, Coronavirus biology and replication: implications for SARS-CoV-2, Nat. Rev. Microbiol. 19 (2021) 155–170. https://doi.org/10.1038/s41579-020-00468-6.

[2] J.-T. Suhren, A. Meinardus, K. Hussein, N. Schaumann, Meta-analysis on COVID-19-pregnancy-related placental pathologies shows no specific pattern, Placenta. 117 (2022) 72–77. https://doi.org/10.1016/j.placenta.2021.10.010.

[3] J.C. Watkins, V.F. Torous, D.J. Roberts, Defining Severe Acute Respiratory Syndrome Coronavirus 2 (SARS-CoV-2) Placentitis, Arch. Pathol. Lab. Med. 145 (2021) 1341–1349. https://doi.org/10.5858/arpa.2021-0246-SA.

[4] M.C. Sharps, D.J.L. Hayes, S. Lee, Z. Zou, C.A. Brady, Y. Almoghrabi, A. Kerby, K.K. Tamber, C.J. Jones, K.M. Adams Waldorf, A.E.P. Heazell, A structured review of placental morphology and histopathological lesions associated with SARS-CoV-2 infection, Placenta. 101 (2020) 13–29. https://doi.org/10.1016/j.placenta.2020.08.018.

[5] M. Mirbeyk, A. Saghazadeh, N. Rezaei, A systematic review of pregnant women with COVID-19 and their neonates, Arch. Gynecol. Obstet. 304 (2021) 5–38. https://doi.org/10.1007/s00404-021-06049-z.

[6] D.D. Flannery, S. Gouma, M.B. Dhudasia, S. Mukhopadhyay, M.R. Pfeifer, E.C. Woodford, J.E. Triebwasser, J.S. Gerber, J.S. Morris, M.E. Weirick, C.M. McAllister, M.J. Bolton, C.P. Arevalo, E.M. Anderson, E.C. Goodwin, S.E. Hensley, K.M. Puopolo, Assessment of Maternal and Neonatal Cord Blood SARS-CoV-2 Antibodies and Placental Transfer Ratios, JAMA Pediatr. 175 (2021) 594–600. https://doi.org/10.1001/jamapediatrics.2021.0038.

[7] A.G. Edlow, J.Z. Li, A.-R.Y. Collier, C. Atyeo, K.E. James, A.A. Boatin, K.J. Gray, E.A. Bordt, L.L. Shook, L.M. Yonker, A. Fasano, K. Diouf, N. Croul, S. Devane, L.J. Yockey, R. Lima, J. Shui, J.D. Matute, P.H. Lerou, B.O. Akinwunmi, A. Schmidt, J. Feldman, B.M. Hauser, T.M. Caradonna, D. De la Flor, P. D’Avino, J. Regan, H. Corry, K. Coxen, J. Fajnzylber, D. Pepin, M.S. Seaman, D.H. Barouch, B.D. Walker, X.G. Yu, A.J. Kaimal, D.J. Roberts, G. Alter, Assessment of Maternal and Neonatal SARS-CoV-2 Viral Load, Transplacental Antibody Transfer, and Placental Pathology in Pregnancies During the COVID-19 Pandemic, JAMA Netw. Open. 3 (2020) e2030455. https://doi.org/10.1001/jamanetworkopen.2020.30455.

[8] N.T. Joseph, C.M. Dude, H.P. Verkerke, L.S. Irby, A.L. Dunlop, R.M. Patel, K.A. Easley, A.K. Smith, S.R. Stowell, D.J. Jamieson, V. Velu, M.L. Badell, Maternal Antibody Response, Neutralizing Potency, and Placental Antibody Transfer After Severe Acute Respiratory Syndrome Coronavirus 2 (SARS-CoV-2) Infection, Obstet. Gynecol. 138 (2021) 189–197. https://doi.org/10.1097/AOG.0000000000004440.

[9] J.C. Marshall, S. Murthy, J. Diaz, N. Adhikari, D.C. Angus, Y.M. Arabi, K. Baillie, M. Bauer, S. Berry, B. Blackwood, M. Bonten, F. Bozza, F. Brunkhorst, A. Cheng, M. Clarke, V.Q. Dat, M. de Jong, J. Denholm, L. Derde, J. Dunning, X. Feng, T. Fletcher, N. Foster, R. Fowler, N. Gobat, C. Gomersall, A. Gordon, T. Glueck, M. Harhay, C. Hodgson, P. Horby, Y.J. Kim, R. Kojan, B. Kumar, J. Laffey, D. Malvey, I. Martin-Loeches, C. McArthur, D. McAuley, S. McBride, S. McGuinness, L. Merson, S. Morpeth, D. Needham, M. Netea, M.D. Oh, S. Phyu, S. Piva, R. Qiu, H. Salisu-Kabara, L. Shi, N. Shimizu, J. Sinclair, S. Tong, A. Turgeon, T. Uyeki, F. van de Veerdonk, S. Webb, P. Williamson, T. Wolf, J. Zhang, A minimal common outcome measure set for COVID-19 clinical research, Lancet Infect. Dis. 20 (2020) e192–e197. https://doi.org/10.1016/S1473-3099(20)30483-7.

[10] World Health Organization, Child growth standards, (n.d.). https://www.who.int/toolkits/child-growth-standards/standards (accessed September 11, 2021).

[11] D. Levitan, V. London, R.A. McLaren, J.D. Mann, K. Cheng, M. Silver, K.S. Balhotra, S. McCalla, K. Loukeris, Histologic and Immunohistochemical Evaluation of 65 Placentas from Women with Polymerase Chain Reaction–Proven Severe Acute Respiratory Syndrome Coronavirus 2 (SARS-CoV-2) Infection, Arch. Pathol. Lab. Med. 145 (2021) 648–656. https://doi.org/10.5858/arpa.2020-0793-SA.

[12] T.Y. Khong, E.E. Mooney, I. Ariel, N.C.M. Balmus, T.K. Boyd, M.A. Brundler, H. Derricott, M.J. Evans, O.M. Faye-Petersen, J.E. Gillan, A.E.P. Heazell, D.S. Heller, S.M. Jacques, S. Keating, P. Kelehan, A. Maes, E.M. McKay, T.K. Morgan, P.G.J. Nikkels, W.T. Parks, R.W. Redline, I. Scheimberg, M.H. Schoots, N.J. Sebire, A. Timmer, G. Turowski, J.P. Van Der Voorn, I. Van Lijnschoten, S.J. Gordijn, Sampling and definitions of placental lesions Amsterdam placental workshop group consensus statement, in: Arch. Pathol. Lab. Med., Arch Pathol Lab Med, 2016: pp. 698–713. https://doi.org/10.5858/arpa.2015-0225-CC.

[13] Y.J. Hou, K. Okuda, C.E. Edwards, D.R. Martinez, T. Asakura, K.H. Dinnon, T. Kato, R.E. Lee, B.L. Yount, T.M. Mascenik, G. Chen, K.N. Olivier, A. Ghio, L.V. Tse, S.R. Leist, L.E. Gralinski, A. Schäfer, H. Dang, R. Gilmore, S. Nakano, L. Sun, M.L. Fulcher, A. Livraghi-Butrico, N.I. Nicely, M. Cameron, C. Cameron, D.J. Kelvin, A. de Silva, D.M. Margolis, A. Markmann, L. Bartelt, R. Zumwalt, F.J. Martinez, S.P. Salvatore, A. Borczuk, P.R. Tata, V. Sontake, A. Kimple, I. Jaspers, W.K. O’Neal, S.H. Randell, R.C. Boucher, R.S. Baric, SARS-CoV-2 Reverse Genetics Reveals a Variable Infection Gradient in the Respiratory Tract, Cell. 182 (2020) 429-446.e14. https://doi.org/10.1016/J.CELL.2020.05.042.

[14] N. Huang, P. Pérez, T. Kato, Y. Mikami, K. Okuda, R.C. Gilmore, C.D. Conde, B. Gasmi, S. Stein, M. Beach, E. Pelayo, J.O. Maldonado, B.A. Lafont, S.I. Jang, N. Nasir, R.J. Padilla, V.A. Murrah, R. Maile, W. Lovell, S.M. Wallet, N.M. Bowman, S.L. Meinig, M.C. Wolfgang, S.N. Choudhury, M. Novotny, B.D. Aevermann, R.H. Scheuermann, G. Cannon, C.W. Anderson, R.E. Lee, J.T. Marchesan, M. Bush, M. Freire, A.J. Kimple, D.L. Herr, J. Rabin, A. Grazioli, S. Das, B.N. French, T. Pranzatelli, J.A. Chiorini, D.E. Kleiner, S. Pittaluga, S.M. Hewitt, P.D. Burbelo, D. Chertow, D.E. Kleiner, M.S. De Melo, E. Dikoglu, S. Desar, K. Ylaya, J.Y. Chung, G. Smith, D.S. Chertow, K.M. Vannella, M. Ramos-Benitez, S.C. Ramelli, S.J. Samet, A.L. Babyak, L.P. Valenica, M.E. Richert, N. Hays, M. Purcell, S. Singireddy, J. Wu, J. Chung, A. Borth, K. Bowers, A. Weichold, D. Tran, R.J. Madathil, E.M. Krause, D.L. Herr, J. Rabin, J.A. Herrold, A. Tabatabai, E. Hochberg, C. Cornachione, A.R. Levine, M.T. McCurdy, K.K. Saharia, Z. Chancer, M.A. Mazzeffi, J.E. Richards, J.W. Eagan, Y. Sangwan, I. Sequeira, S. A. Teichmann, A. J. Kimple, K. Frank, J. Lee, R.C. Boucher, S.A. Teichmann, B.M. Warner, K.M. Byrd, SARS-CoV-2 infection of the oral cavity and saliva, Nat. Med. 27 (2021) 892–903. https://doi.org/10.1038/S41591-021-01296-8.

[15] North Carolina’s Hispanic Community: 2019 Snapshot | Carolina Demography, (n.d.). https://www.ncdemography.org/2019/09/26/north-carolinas-hispanic-community-2019-snapshot/ (accessed September 11, 2021).

[16] P.Z. Rebutini, A.C. Zanchettin, E.T.S. Stonoga, D.M.M. Prá, A.L.P. de Oliveira, F. da S. Dezidério, A.S. Fonseca, J.C.H. Dagostini, E.C. Hlatchuk, I.N. Furuie, J. da S. Longo, B.M. Cavalli, C.L.T. Dino, V.M. de C.H. Dias, A.P. Percicote, M.B. Nogueira, S.M. Raboni, N.S. de Carvalho, C. Machado-Souza, L. de Noronha, Association Between COVID-19 Pregnant Women Symptoms Severity and Placental Morphologic Features, Front. Immunol. 12 (2021). https://doi.org/10.3389/fimmu.2021.685919.

[17] D.A. Schwartz, An analysis of 38 pregnant women with COVID-19, their newborn infants, and maternal-fetal transmission of SARS-CoV-2: Maternal coronavirus infections and pregnancy outcomes, Arch. Pathol. Lab. Med. 144 (2020) 799–805. https://doi.org/10.5858/arpa.2020-0901-SA.

[18] F. Parazzini, R. Bortolus, P.A. Mauri, A. Favilli, S. Gerli, E. Ferrazzi, Delivery in pregnant women infected with SARS-CoV-2: A fast review, Int. J. Gynecol. Obstet. 150 (2020) 41–46. https://doi.org/10.1002/ijgo.13166.

[19] B.J.F. Huntley, E.S. Huntley, D. Di Mascio, T. Chen, V. Berghella, S.P. Chauhan, Rates of maternal and perinatal mortality and vertical transmission in pregnancies complicated by severe acute respiratory syndrome coronavirus 2 (SARS-Co-V-2) infection: A systematic review, Obstet. Gynecol. 136 (2020) 303–312. https://doi.org/10.1097/AOG.0000000000004010.

[20] J. Juan, M.M. Gil, Z. Rong, Y. Zhang, H. Yang, L.C. Poon, Effect of coronavirus disease 2019 (COVID-19) on maternal, perinatal and neonatal outcome: systematic review, Ultrasound Obstet. Gynecol. 56 (2020) 15–27. https://doi.org/10.1002/uog.22088.

[21] D. Di Mascio, A. Khalil, G. Saccone, G. Rizzo, D. Buca, M. Liberati, J. Vecchiet, L. Nappi, G. Scambia, V. Berghella, F. D’Antonio, Outcome of coronavirus spectrum infections (SARS, MERS, COVID-19) during pregnancy: a systematic review and meta-analysis, Am. J. Obstet. Gynecol. MFM. 2 (2020) 100107. https://doi.org/10.1016/j.ajogmf.2020.100107.

[22] K.F. Walker, K. O’Donoghue, N. Grace, J. Dorling, J.L. Comeau, W. Li, J.G. Thornton, Maternal transmission of SARS-COV-2 to the neonate, and possible routes for such transmission: a systematic review and critical analysis, BJOG Int. J. Obstet. Gynaecol. 127 (2020) 1324–1336. https://doi.org/10.1111/1471-0528.16362.

[23] M. Guan, E. Johannesen, C.Y. Tang, A.L. Hsu, C.L. Barnes, M. Burnam, J.A. McElroy, X.-F. Wan, Intrauterine fetal demise in the third trimester of pregnancy associated with mild infection with the SARS-CoV-2 Delta variant without protection from vaccination, J. Infect. Dis. (2022) jiac007. https://doi.org/10.1093/infdis/jiac007.

[24] Stats of the States - Cesarean Delivery Rates, (n.d.). https://www.cdc.gov/nchs/pressroom/sosmap/cesarean_births/cesareans.htm (accessed September 11, 2021).

[25] T. Boyd, D. Gang, G. Lis, A. Juozokas, S. Pflueger, Normative values for placental weights (N= 15,463), Mod Pathol. 12 (1999).

