## Supplementary figures and images for "The clinical impact of maternal COVID-19 on mothers, their infants, and placentas with an analysis of vertical transfer of maternal SARS-CoV-2-specific IgG antibodies"

### Supplemental Figure 1

Survival of Infants Born To Mothers With COVID19: By Trimester of Infection

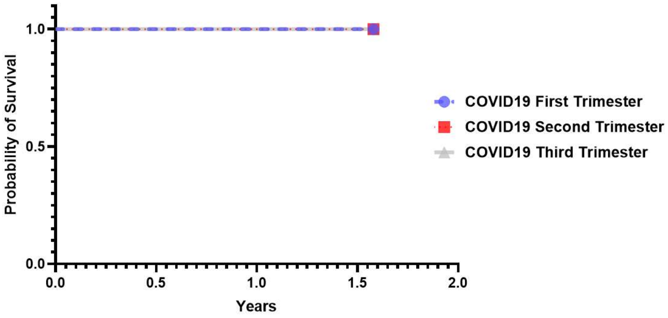

### Supplemental Figure 2

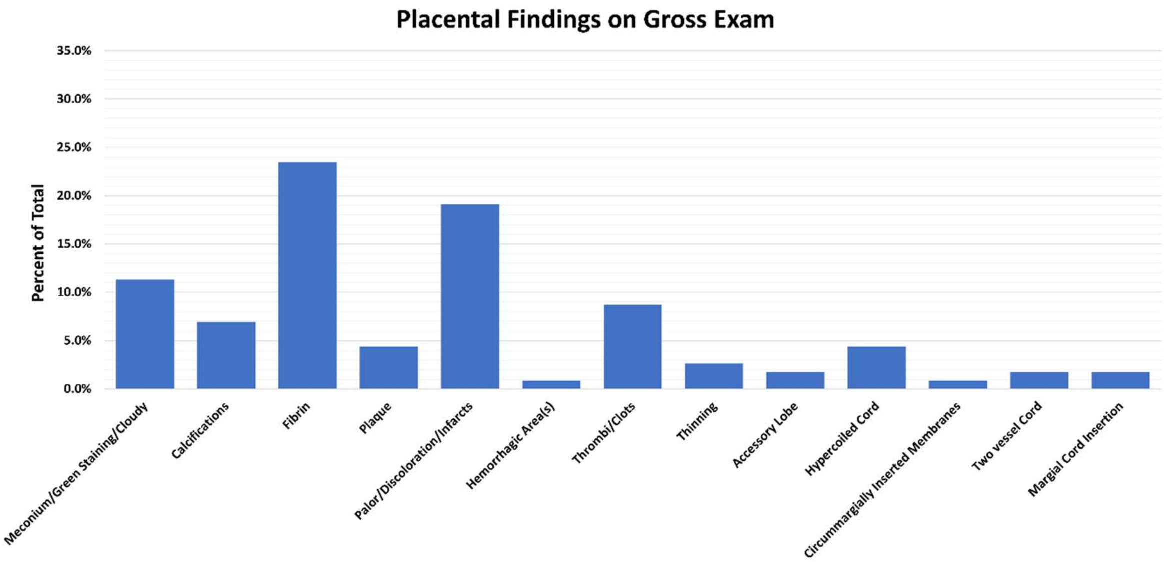

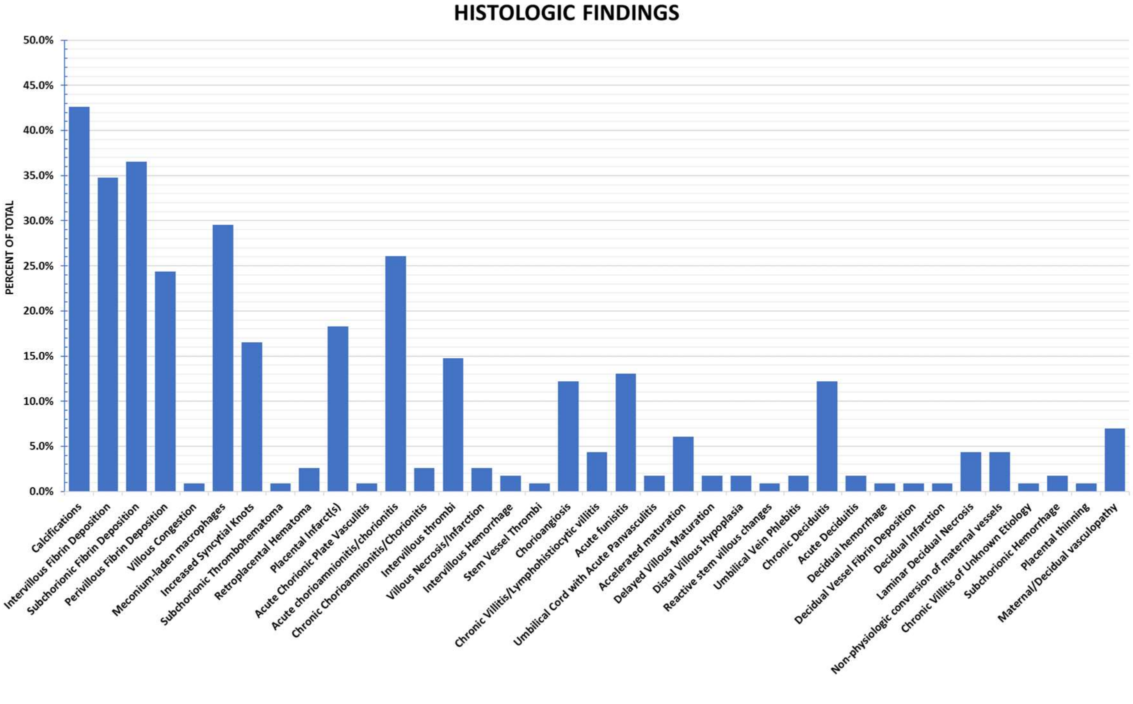

### Supplemental Figure 3

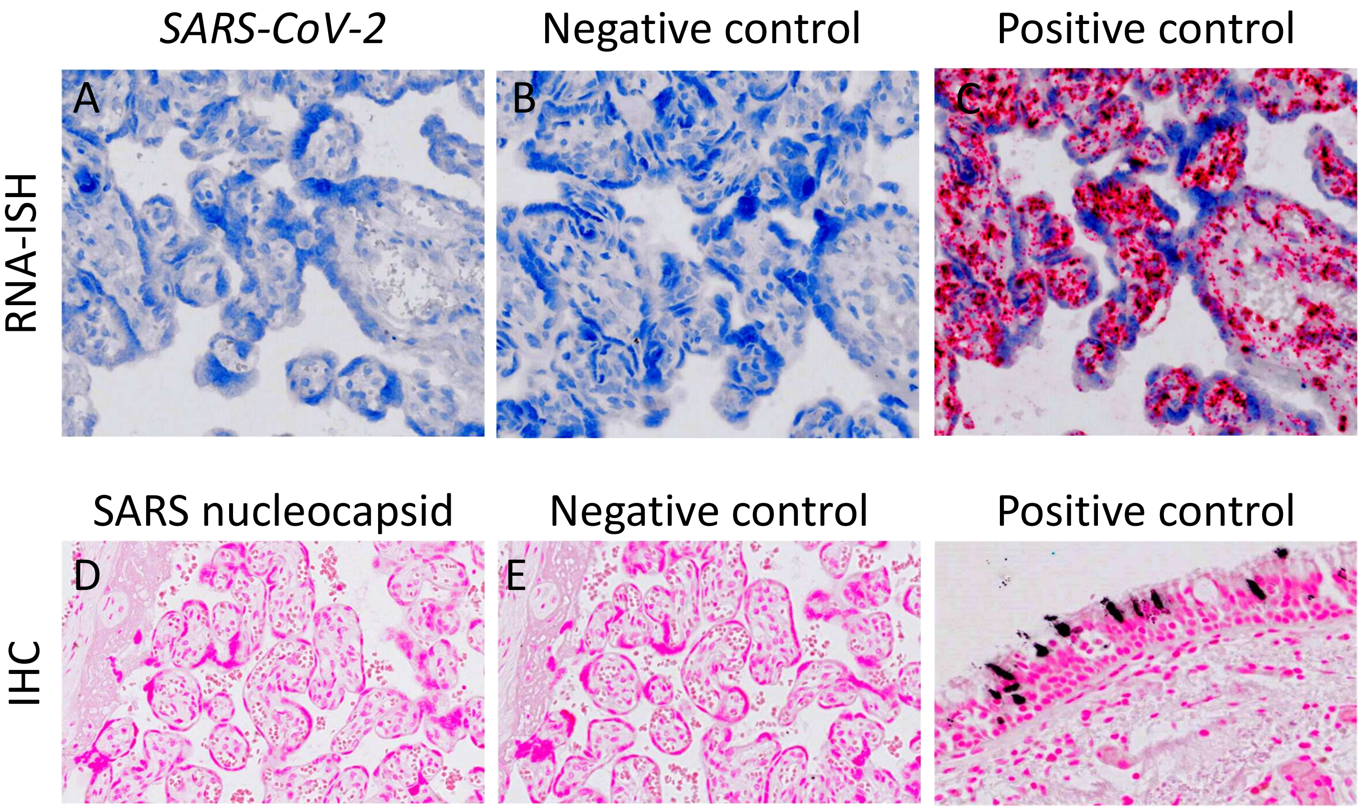

### Supplemental Figure 4

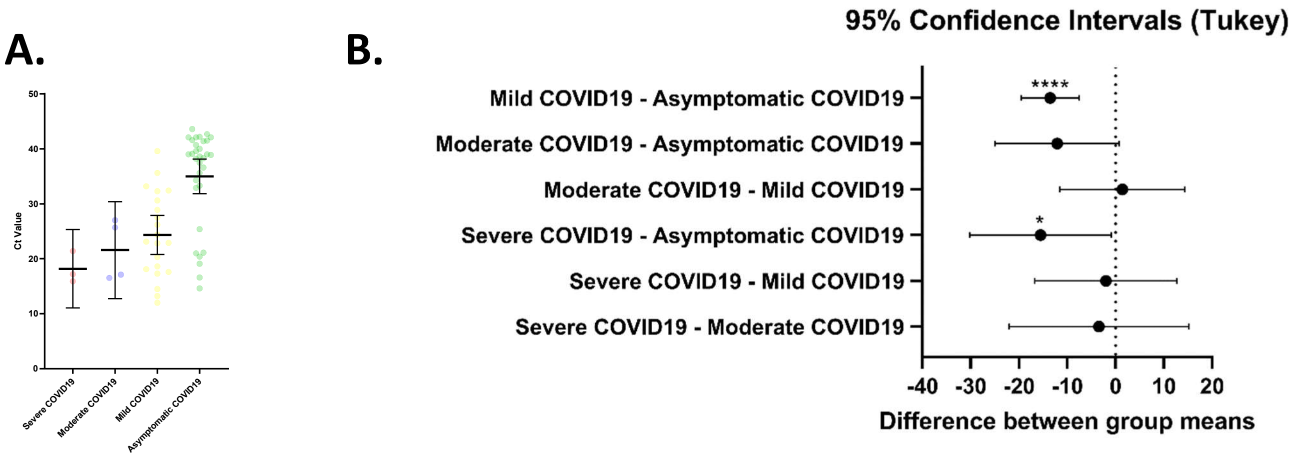
