## Supplemental Table 1 and Supplement Figure Legends for "The clinical impact of maternal COVID-19 on mothers, their infants, and placentas with an analysis of vertical transfer of maternal SARS-CoV-2-specific IgG antibodies"

**Supplemental Table 1: Detailed results of SARS-CoV-2-neutralizing IgG mother and infant cohort**

| **Sample** | **Avg % Inhibition Value** | **Avg Neut Ab Conc (ng/ml)** |
| --- | --- | --- |
| Mother 1 | 49.52 | 112.60 |
| Baby 1 | 54.85 | 131.54 |
| Mother 2 | 62.87 | 167.82 |
| Baby 2 | 70.73 | 218.65 |
| Mother 3 | 58.27 | 145.64 |
| Baby 3 | 59.60 | 151.66 |
| Mother 4 | 18.25 | **below LOQ** |
| Baby 4 | 32.11 | 66.47 |
| Mother 5 | 55.00 | 132.15 |
| Baby 5 | 70.65 | 218.02 |
| Mother 6 | 77.64 | 213.97 |
| Baby 6 | 84.04 | 300.09 |
| Mother 7 | 68.70 | 147.51 |
| Baby 7 | 78.20 | 207.09 |
| Mother 8 | 0.14 | **below LOQ** |
| Baby 8 | 0.00 | **below LOQ** |
| Mother 9 | 25.85 | 38.13 |
| Baby 9 | 3.45 | **below LOQ** |
| Mother 10 | 43.54 | 55.91 |
| Baby 10 | 57.96 | 99.11 |
| Pos Control | 97.35 |  |
| Neg Control | 13.86 |  |

LOQ = Limit of quantitation

**Supplemental Figure 1: Survival curves of infants born to mothers with SARS-CoV-2 by trimester of maternal SARS-CoV-2 infection.**

**Supplemental Figure 2: Incidence of specific gross and microscopic findings of evaluated placentas.** n=115.

**Supplemental Figure 3: RNAScope for SARS-CoV-2 and immunohistochemistry for SARS-CoV-2 spike protein demonstrate the absence of placental infection with SARS-CoV-2 in the single SARS-CoV-2 positive infant in the SARS-CoV-2-specific IgG antibody test cohort.** As with the placenta of the single SARS-CoV-2 positive infant in our consecutive cohort, the placenta of the single SARS-CoV2 positive infant in the serum study cohort demonstrated a lack of SARS-CoV-2 RNA or the presence of SARS-CoV-2 spike protein, indicating that the infant’s SARS-CoV-2 infection was not due to vertical transmission of SARS-CoV-2.

**Supplemental Figure 4: Comparison of cycle threshold (Ct) values of SARS-CoV-2 amplification based on COVID-19 severity. A:** Box and whiskers plot of Ct values among categories of COVID-19 severity. Bar represents mean with error bars representing 95% confidence interval. **B:** One-way ANOVA with Tukey’s multiple comparison test. Error bars represent 95% confidence intervals. *p=0.0339, ****p<0.0001
